# Effectiveness of Therapies Treating Sensory Processing Disorders in Children with Autism Spectrum Disorder

**DOI:** 10.1101/2024.09.17.24313823

**Authors:** Joshua Orgel

**Affiliations:** Rowan Virtua School of Osteopathic Medicine Medical Scholarship Two

## Abstract

The prevalence of children being diagnosed with Autism Spectrum Disorder has risen dramatically over recent years. Many of those children suffer from sensory processing disorders (SPD). There is no standardized treatment for this disorder and the therapies available have produced confusing and conflicting results. A search was performed for relevant studies and systematic review articles that researched treatment in this area. Data was extracted and analyzed and then organized with summary conclusions of each paper. Qualitative notes were gathered on the studies and the results of the studies were compared and contrasted. Sensory integration therapy has been shown to produce significant results and most of the conflicting data can be explained scientifically. Research is still required for the duration frequency and rate of therapy. Reviews of other therapies to treat SPD have not shown enough evidence of significant positive outcomes to be recommended.

**Purpose Statement:** The purpose of this paper is to gather and analyze research regarding therapies for Sensory Processing Disorder in Children with Autism Spectrum Disorder. This review aims to determine the evidence of such therapies and to inform and educate practitioners regarding outcomes in their use to treat this disorder.

## Introduction

The prevalence of Autism Spectrum Disorder (ASD) has grown increasingly within recent years. In the year 2000 there were 6.7 per 1000 children diagnosed with ASD and in the year 2018 there were 23 children per 1000 children diagnosed. It is approximately 4 times more common in males than females.^1^ ASD is a neurological disorder that is mainly diagnosed in the early stage of life. No single cause or biomarker has been identified and there are broad range symptoms within the spectrum. Although the symptoms are varied, ASD is categorized by pervasive difficulties with communication and social interaction with others, restrictive interests, restrictive or repetitive behaviors, and symptoms that affect school work and home settings.^2^

It has been established that one of the symptoms that affect many of those with ASD is sensory processing difficulty. In the DSM V, abnormal reactions to sensory stimuli is listed as one of the diagnostic criterion of ASD.^3^ In one study the abnormal response to sensory stimuli in the first two years of life separated children who were diagnosed with ASD from those who suffered from a learning disability or who had a lower IQ.^4^ Sensory symptoms are predictive of social communication deficits and repetitive behaviors in childhood.^5^ An estimated 80% of children diagnosed with autism display sensory processing difficulties.^6^ These sensory processing difficulties often lead to negative behaviors and decreased functioning within the child’s environment. Hyperreactivity to stimuli such as loud sounds, touch, or overwhelming environment may lead to distress, avoidance, or hypervigilance. Children who respond to stimuli in a hyporeactive manner may appear unaware or non-responsive to the stimuli or they may actively seek new intense stimuli to increase their arousal.^3^ Children with sensory processing difficulties may actively seek or avoid ordinary sensations such as auditory, tactile, or vestibular input.^3^

Currently there is no known treatment for Autism and the available treatment is directed towards decreasing symptoms and increasing functioning within daily activities.^1^ Although sensory interventions are common for ASD children a broad range of treatments are implemented with conflicting evidence of efficacy.^7^ Sensory-based intervention is one of the most common therapies requested from parents for children with ASD.^3^ Some of the more mainstream treatments include Sensory Integration training (SIT) and Sensory Based Interventions, (SBI). Less common treatments include music therapy, massage therapy, and auditory integration training (AIT). Research within this field has been tumultuous. Since there is mostly no standard or set protocol even with one given type of treatment any research has been hard to compare or even reproduce. Additionally, there is no broadly agreed-upon standard regarding the assessment of a child’s sensory processing difficulties. This creates difficulties regarding scoring a sensory profile before and after a researched treatment. Some of the research has been performed by professionals other than those in the medical field and terminology can be confusing when comparing and analyzing research.

## Methods

### Search Term Strategy

On December 1 2022 we identified potential studies by searching PUBMED for peer-reviewed articles using keywords. The following search terms were used: “Autism” OR “ASD” AND “sensory” AND “therapy”, OR “Intervention” OR “integration” OR “sensory processing”.

### Inclusion Criteria

Any articles that contained original research of treatment targeting sensory processing difficulties with children diagnosed with autism or research that reviewed papers that included such were included. Original research included clinical studies and retrospective studies and review papers included systematic reviews and meta-analysis papers. Foreign papers that were translated in English were also included. Case studies were not included.

### Participants

Only pediatric participants were included. A pediatric was defined as age 0 to 18. Both males and females were included, and all genders, races, and nationalities were included.

### Types of treatments

Only research that included treatment with sensory integration principles or sensory-based intervention or music therapy or massage therapy or auditory integration therapy were included. There is very limited research regarding other treatments when using the above quoted search terms.

### Data extraction

Article and research information was extracted from the papers that were produced using the above search terms and that fit into the inclusion criteria. Information was taken on the names of the authors, the date the article was published and the country of the journal it was published. Data from original research including sample size, types of assessments and treatments used, outcomes, and quantitative and qualitative analysis were extracted.

### Data analysis

Extracted data will be gathered, summarized and analyzed descriptively. A review of the efficacy of the various treatments for Children with ASD and Sensory processing difficulties was conducted.

## Results

A total of 7 clinical studies of Sensory integration therapy were identified. Studies included 3 randomized control studies, 3 cohort studies, and one pilot study. Studies that used the term ASI or Aryes sensory integration were included together with studies of SIT or sensory integration therapy. A total of 6 studies identified positive outcomes in at least one area. One study found that there were no significant differences in outcomes between children treated with sensory integration therapy and those treated with usual care. All studies showed improvement in motor skills however, there were varied results regarding other measured goals which showed varied outcomes.

**Table 1.** Organization of clinical trials researched treatment of sensory processing difficulty in ASD children with Sensory Integration Therapy. Includes authors of the papers, date published, participants, type of therapy used, notes from the conclusion of the papers and additional notes from the author of this paper. The papers were arranged in chronological published date order.

| STI Clinical | Year | Type of Research | Number of Subjects/<br>Studies Reviewed | Type of Treatment/therapy applied/<br>reviewed | Conclusion statement | Additional notes |
| --- | --- | --- | --- | --- | --- | --- |
| Case-Smith J, Bryan T.<sup>8</sup> | 1999 | Cohort study | 5 participants | SIT | 3/5 significant improvement in mastery play. 4/5 showed less non engaged play. Non direct targeted measures did not seem to improve significantly | 10 weeks<br>Did not directly measure sensory processing<br>Small sample size |
| Pfeiffer BA, Koenig K, Kinnealey M, Sheppard M, Henderson L.<sup>9</sup> | 2011 | Pilot study (SIT vs Fine motor intervention) | 33 participants | SIT | Children in the SIT group had significantly fewer autistic mannerisms after interventions.<br>No significant differences were found in the scores on the SPM (sensory processing measure) or the QNST-II (neurological interference) | Researchers noted that due to nature of study treatment and evaluation in clinic. Need for carryover to home which could have affected SPM score (which was partially scored by parents) |
| Iwanaga R,<br>Honda S,<br>Nakane H,<br>Tanaka K,<br>Toeda H,<br>Tanaka G.<sup>10</sup> | 2014 | cohort | 8 participants | SIT | The changes in score from before and after therapy indicated that SIT might improve fundamental sensory-motor abilities, coordination abilities, non verbal cognitive abilities, and visual-motor abilities | Small sample size<br>Used Japanese version of assessment which there has been so significant data regarding reliability. |
| Karim AEA, Mohammed AH<sup>11</sup> | 2015 | Cohort study | 34 participants | SIT | The results of this study showed significant improvement in their motor skills after receiving sensory integration therapy. | Only reviewed SIT regarding motor improvement<br>3 session/week over 6 months |
| Kashefi mehr B, Kayihan H, Huri M<sup>12</sup> | 2018 | Randomized Control Study<br>(Treatment vs waiting list) | 35 participants | SIT/SBI | significantly greater improvement was observed in the intervention group in all domains of SCOPE including volition, habituation, communication and interaction skills, process skills, motor skills, and environment of occupational performance | 2x week over 12 weeks<br>45 min SIT and 15 min parent training (SBI training) |
| Xu W, Yao J, Liu W.<sup>13</sup> | 2019 | Randomized clinical trial<br>(SIT with education and psychotherapy vs education and psychotherapy without SIT) | 103 participants | SIT | Compared with routine treatment, SIT is better in improving autistic symptoms in children with autism | SIT was used in conjunction with “routine care”<br>Did not state rate or duration of treatment |
| Randell E, Wright M, Milosevic S, et al<sup>14</sup> | 2022 | Randomized Control Study<br>(Vs usual care) | 138 participants | SIT (called ASI or Aryes sensory integration within the article) | The intervention did not demonstrate clinical benefit above standard care | Most recent study<br>26 one hour sessions. 2x week – 10 weeks and then 2x month 2 months<br>Included 2x month phone call<br>Level 1 clinical trial |

**Table 2.**
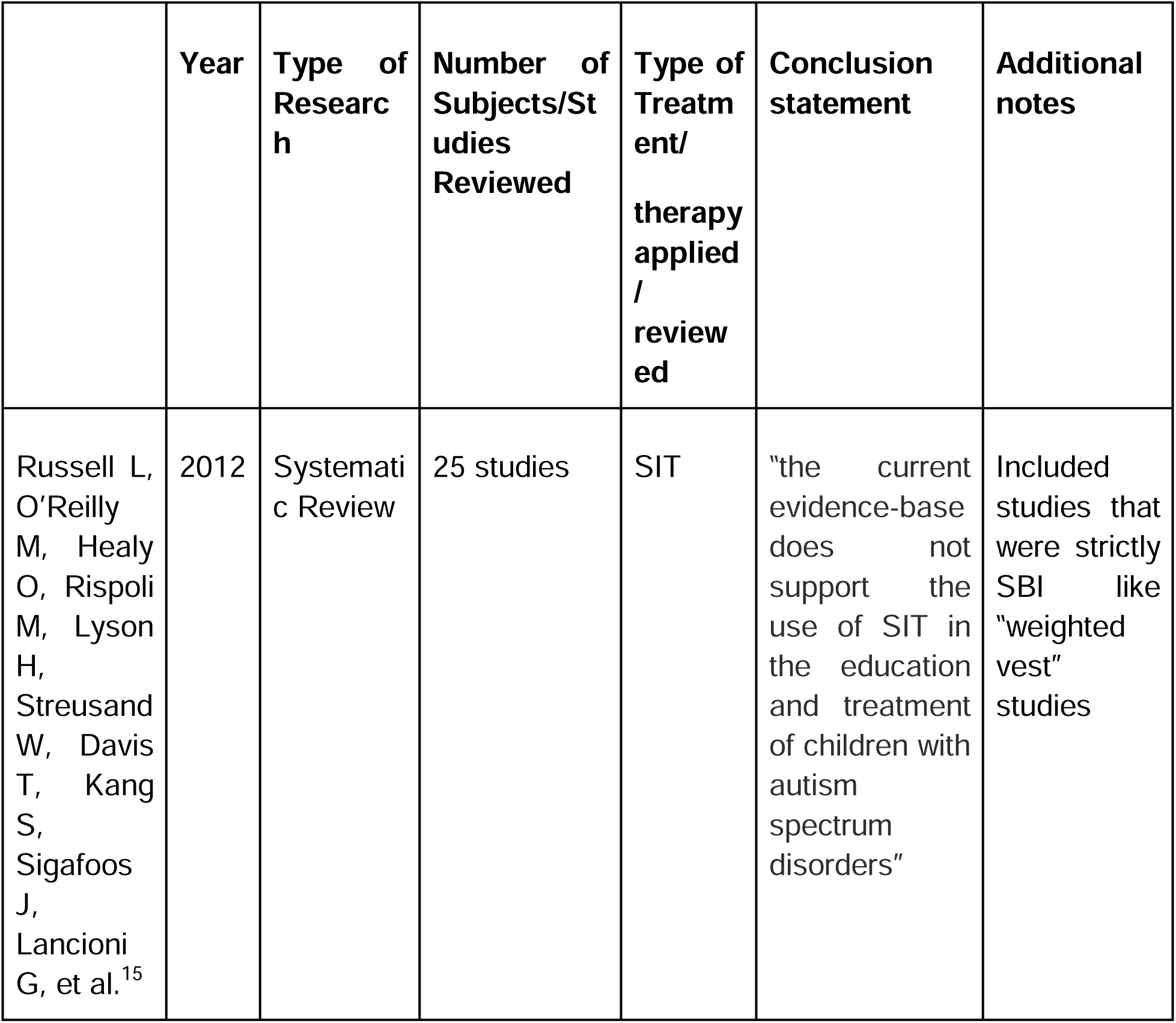
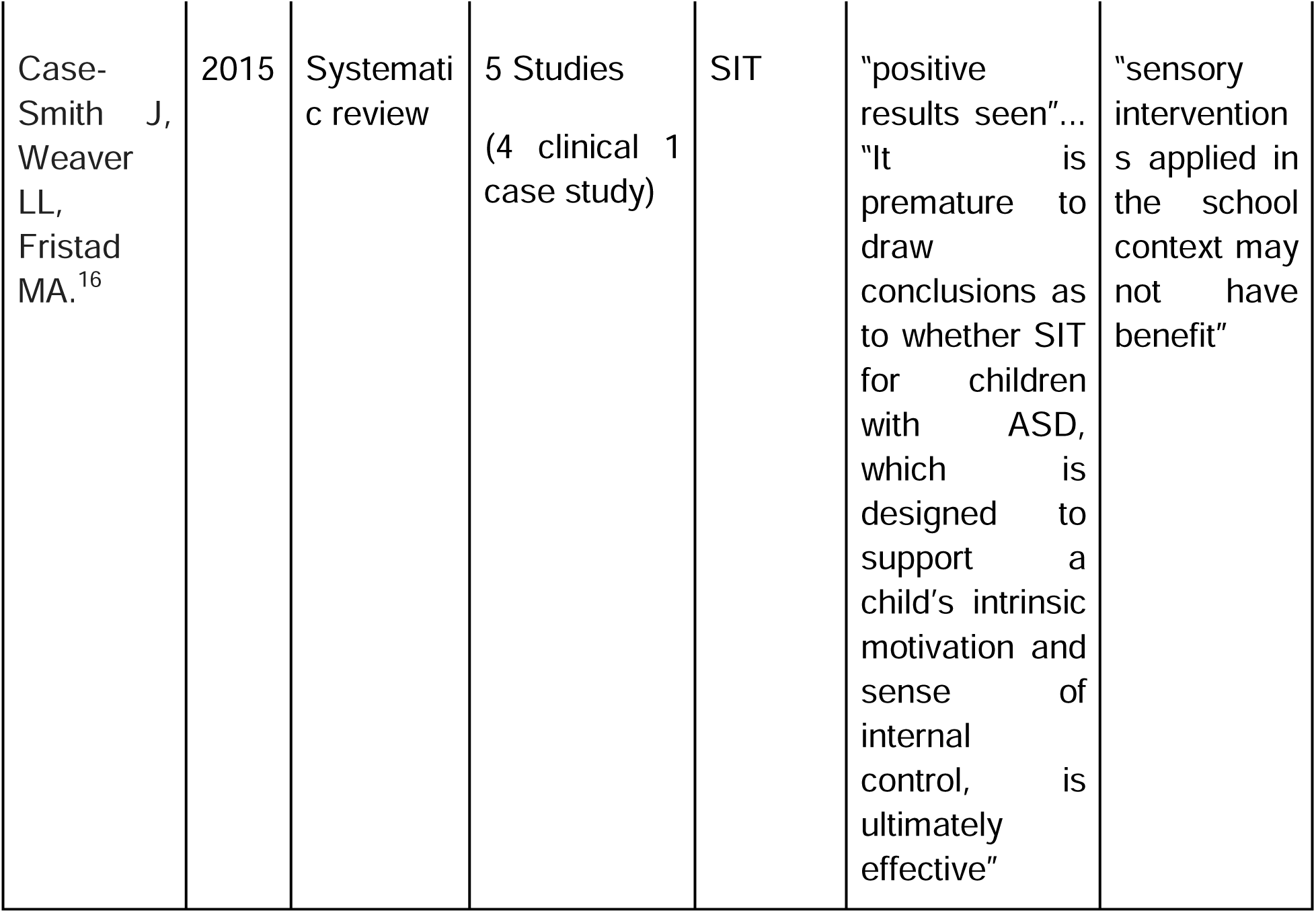
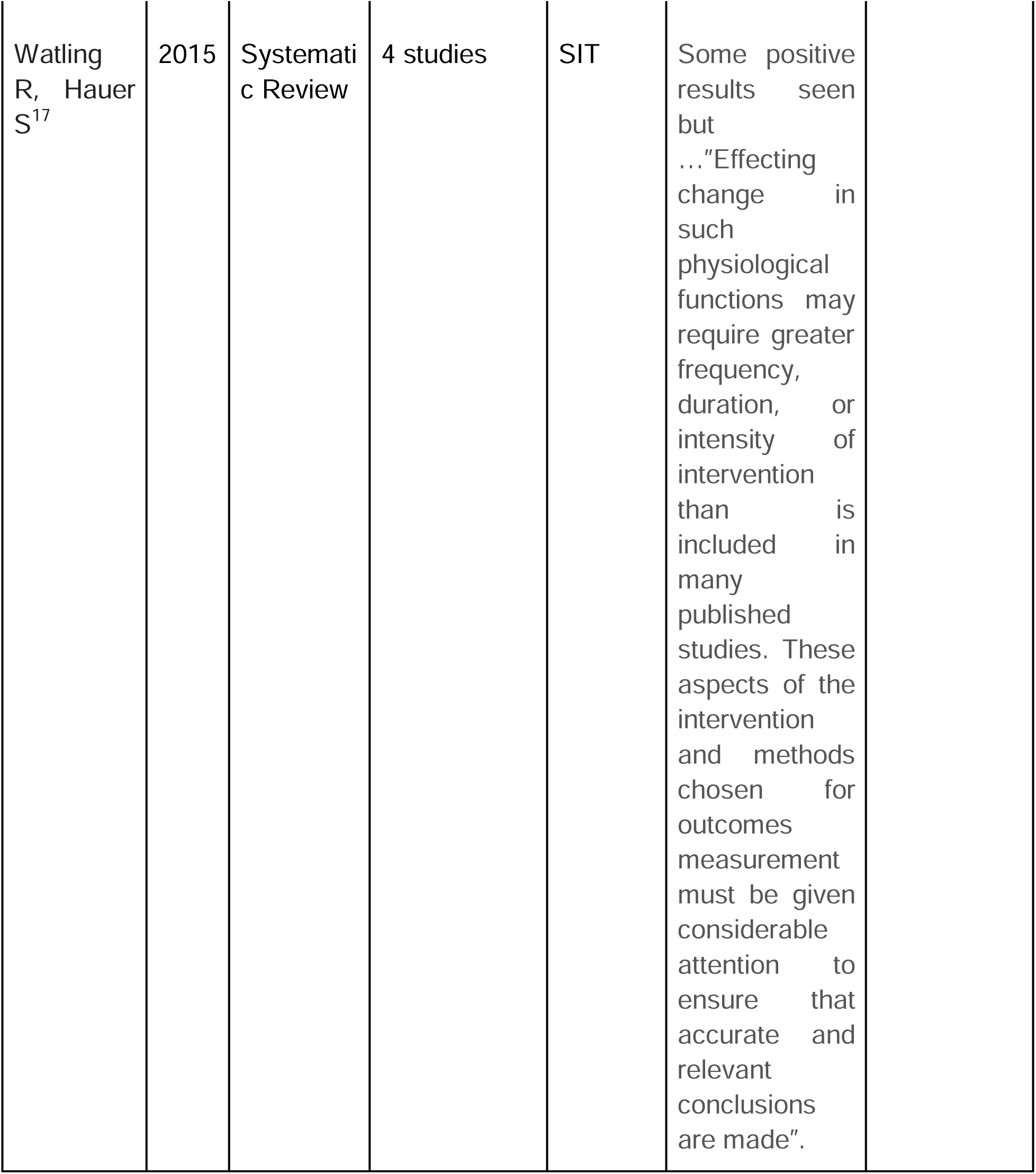
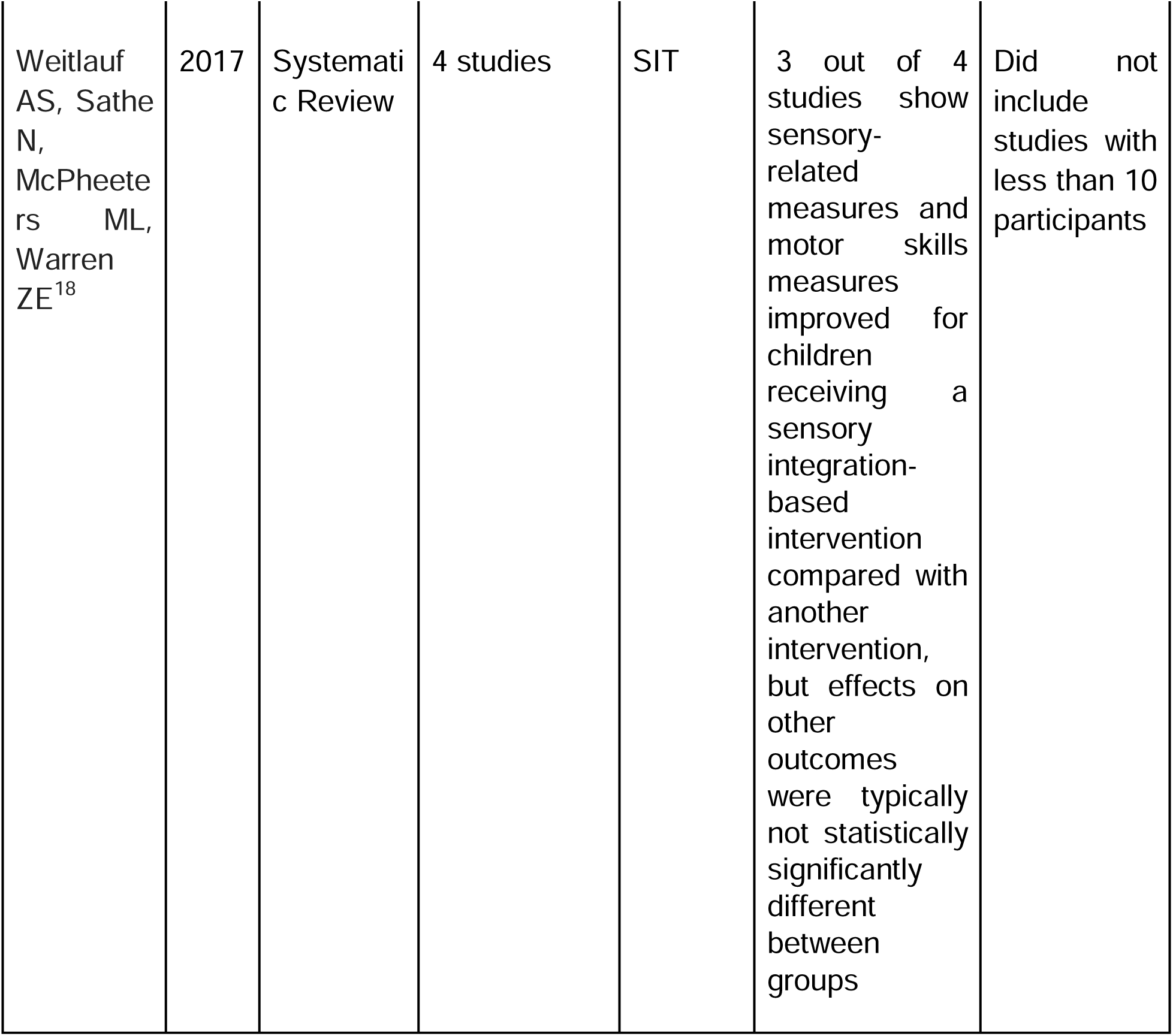
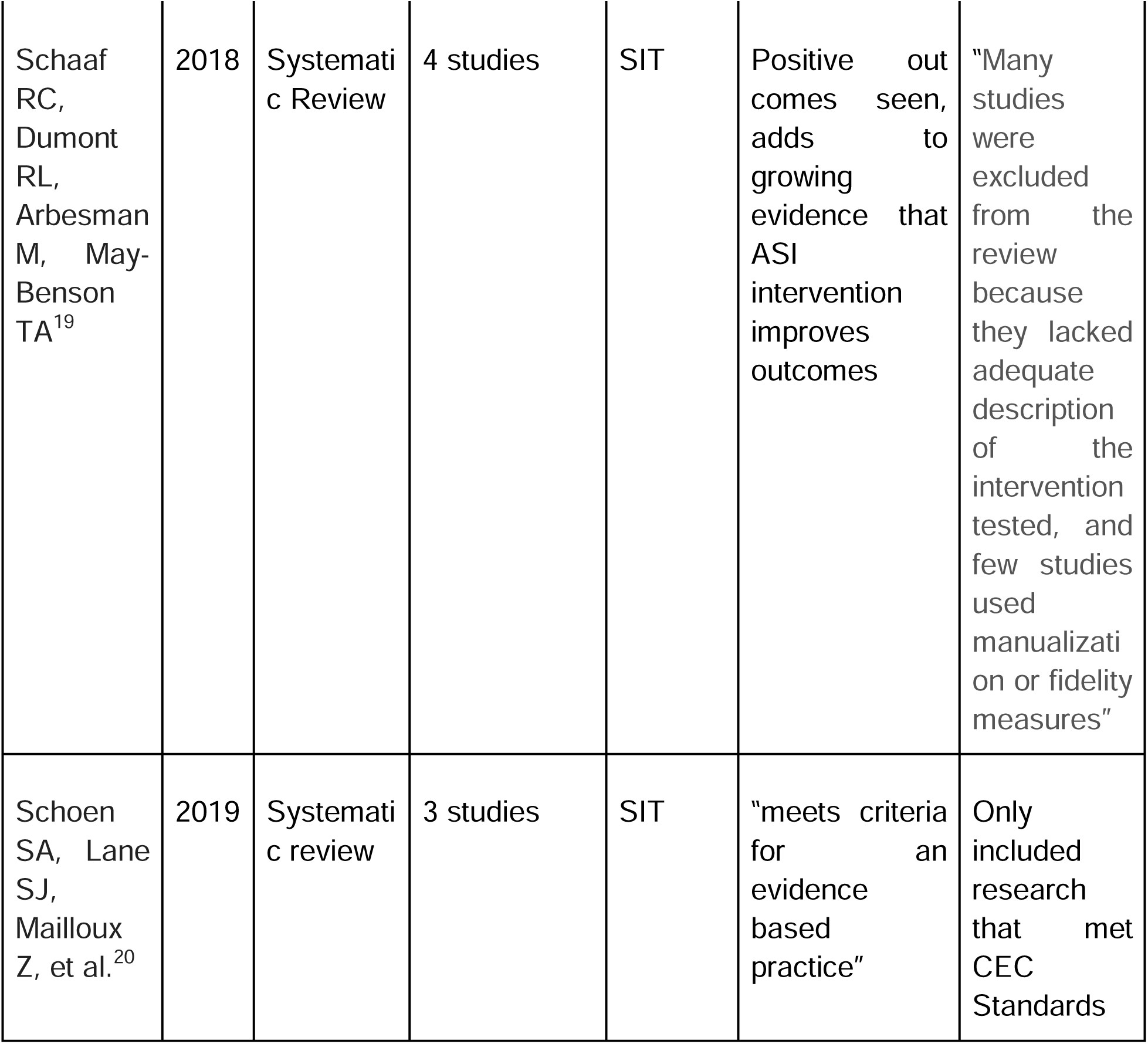
Organization of review papers covering sensory processing difficulty in ASD children with Sensory Integration Therapy. Includes authors of the papers, date published, participants, type of therapy used, notes from the conclusion of the papers and additional notes from the author of this paper. The papers were ordered in chronological published date order.

|  | **Year** | **Type of Research** | **Number of Subjects/Studies Reviewed** | **Type of Treatment/therapy applied / reviewed** | **Conclusion statement** | **Additional notes** |
| --- | --- | --- | --- | --- | --- | --- |
| Russell L, O'Reilly M, Healy O, Rispoli M, Lyson H, Streusand W, Davis T, Kang S, Sigafoos J, Lancioni G, et al.<sup>15</sup> | 2012 | Systematic Review | 25 studies | SIT | "the current evidence-base does not support the use of SIT in the education and treatment of children with autism spectrum disorders" | Included studies that were strictly SBI like "weighted vest" studies |
| Case-Smith J,<br>Weaver LL,<br>Fristad MA.<sup>16</sup> | 2015 | Systematic review | 5 Studies<br>(4 clinical 1 case study) | SIT | “positive results seen”...<br>“It is premature to draw conclusions as to whether SIT for children with ASD, which is designed to support a child’s intrinsic motivation and sense of internal control, is ultimately effective” | “sensory interventions applied in the school context may not have benefit” |

|  |  |  |  |  |  |
| --- | --- | --- | --- | --- | --- |
| Watling<br>R, Hauer<br>S<sup>17</sup> | 2015 | Systematic Review | 4 studies | SIT | Some positive results seen but ...”Effecting change in such physiological functions may require greater frequency, duration, or intensity of intervention than is included in many published studies. These aspects of the intervention and methods chosen for outcomes measurement must be given considerable attention to ensure that accurate and relevant conclusions are made”. |

**Table 2.** Organization of review papers covering sensory processing difficulty in ASD children with Sensory Integration Therapy.
|  |  |  |  |  |  |  |
| --- | --- | --- | --- | --- | --- | --- |
| Weitlauf AS, Sathe N, McPheeters ML, Warren ZE<sup>18</sup> | 2017 | Systematic Review | 4 studies | SIT | 3 out of 4 studies show sensory-related measures and motor skills measures improved for children receiving a sensory integration-based intervention compared with another intervention, but effects on other outcomes were typically not statistically significantly different between groups | Did not include studies with less than 10 participants |
| Schaaf RC, Dumont RL, Arbesman M, May-Benson TA<sup>19</sup> | 2018 | Systematic Review | 4 studies | SIT | Positive outcomes seen, adds to growing evidence that ASI intervention improves outcomes | “Many studies were excluded from the review because they lacked adequate description of the intervention tested, and few studies used manualization or fidelity measures” |
| Schoen SA, Lane SJ, Mailloux Z, et al.<sup>20</sup> | 2019 | Systematic review | 3 studies | SIT | “meets criteria for an evidence based practice” | Only included research that met CEC Standards |

**Table 3.** Organization of review papers and original research of sensory processing difficulty in ASD children with therapies other than Sensory Integration Therapy. Includes authors of the papers, date published, participants, type of therapy used, notes from the conclusion of the papers and additional notes from the author of this paper. The papers were arranged in chronological published date order.

| **Other Studies** | **Year** | **Type of Research** | **Number of Subjects/Studies Reviewed** | **Type of Treatment / therapy applied/ reviewed** | **Conclusion statement** | **Additional notes** |
| --- | --- | --- | --- | --- | --- | --- |
| Weitlauf AS,<br>Sathe N,<br>McPheeters ML,<br>Warren ZE<sup>18</sup> | 2017 | Systematic Review | 2 studies reviewed | Environment based (SBI) | Decrease seen in ASD type behavior and improvement seen in non verbal cognitive skills | Authors dismissed improvement as children still met cut off for ASD even after treatment |
| Case-Smith J,<br>Weaver LL,<br>Fristad MA.<sup>21</sup> | 2015 | Systematic review | 14 studies | SBI | "Outcomes provide very limited evidence of positive effects" |  |
| Watling R, Hauer S<sup>17</sup> | 2015 | Systematic Review | 5 studies multi sensory<br>7 studies single sensory | SBI | Multi-sensory SBIS showed improvement in ASD symptoms. Studies of single-sensory SBIs found very small effects or no effects. | No rigorous level 1 trials have proven that not effective either. |
| Al-Ayadhi L, Al-Drees AM, Al-Arfaj AM<sup>22</sup> | 2013 | Prospective cohort Study | 72 subjects | AIT – Auditory Integration Therapy | All subjects showed significant improvement across 3 different scales used. CARS, SRS and ATEC. Improvement in social awareness, social cognition and social communication. |  |
| Weitlauf AS, Sathe N, McPheeters ML, Warren ZE<sup>18</sup> | 2017 | Systematic Review | 2 studies | AIT – Auditory Integration Therapy | No evidence of effectiveness. | The 2 studies were reported in same publication. |
| Weitlauf AS,<br>Sathe N,<br>McPheeters ML,<br>Warren ZE<sup>18</sup> | 2017 | Systematic Review | 5 studies | Massage therapy | “Massage improved sensory challenges and ASD symptom severity compared with no massage. Our confidence in this conclusion is low (low SOE). Massage did not improve maladaptive behavior (low SOE).” |  |
| Weitlauf AS,<br>Sathe N,<br>McPheeters ML,<br>Warren ZE<sup>18</sup> | 2017 | Systematic Review | 5 studies | Music Therapy | No significant group outcome measures between treated groups and non treated groups | 1 research paper did show improvement |

Six systemic reviews of Sensory integration therapy for children diagnosed with ASD were identified. Of the six, five studies concluded that positive results were seen and one study stating that there was not enough evidence to suggest that STI provides significant positive outcomes. However, one study reported positive results were limited to motor difficulties and that other areas including sensory difficulties and ASD-like behaviors did not have significant outcomes. Two other studies reported positive outcomes but both stated more research was needed in this area to confirm the results and provide further details like rate, duration and frequency.

There were 7 other studies of other therapies besides SIT. Three papers reviewed Sensory Based Intervention (SBI). All studies included isolated sensory-specific strategies that were grouped under SBI. Examples include asking the child to wear a weighted vest, brushing the skin with a specialized sensory brush, use of a therapy ball. One study showed some positive outcomes with SBI but concluded that the improvement was significant enough since the children only moved one level within the ASD criteria and had still met the cut-off for an ASD diagnosis. Another study stated that their review provided little evidence of positive outcomes. The last study split SBI into two groups, Single sensory approach and multi sensory approach. Single sensory was defined as use of a single technique i.e weighted vest and multi sensory approach defined as use of multiple treatments in place. The review found that a single sensory approach provided little evidence of positive outcomes like the previous study. However, the multi-sensory SBI was found to produce a significant decrease in ASD symptoms. Two systematic studies reviewed Auditory Integration Therapy (AIT) to treat sensory processing disorder (SPD) in children with ASD; one review found significant improvement across broad areas while the other review found no evidence of effectiveness. One study reviewed massage therapy to treat SPD in ASD and found there to be significant positive outcomes, The last study reviewed music therapy and found no significant difference between ASD children treated with music therapy and those not treated.

## Discussion

Treating SPD in children with autism is not straightforward. There are many therapies and all of them have different degrees of conflicting evidence if they produced positive outcomes. Many of the first studies done had limited number of participants and were run as cohort studies and not as randomized clinical trials. In 2012 studies were still lumping SBI together with SIT as seen in Russel L (2012) where the authors stated “Implementation of SIT typically involves some combination of the child wearing a weighted vest, being brushed or rubbed with various instruments, riding a scooter board, swinging, sitting on a bouncy ball, being squeezed between exercise pads or pillows, and other similar activities.”^15^ It was not until 2015 that SIT became discrete therapies from SBI as seen in Case-Smith (2015) and Watling (2015)^17^ Of the 4 articles that reviewed SIT only one stated that were was no evidence to suggest positive outcomes, the Russel L15. As mentioned earlier, one study stated there was a positive outcome in SBI with the multi sensory approach, but all agreed that single sensory approach did not about significant positive outcomes. The remaining 3 studies that focused solely on SIT showed positive outcomes and one can say there is no conflicting evidence among review papers regarding this.

Of the 7 clinical trials reviewed the first 4 did not provide high confidence levels of evidence within their outcomes. Two had less than 10 participants, another was a pilot study and another only measured outcomes when it came to motor deficits within children, not ASD mannerisms, behaviors or the child’s sensory processing difficulty.

Adding to the confluence of confusion is that within SIT there have been no set of fidelity measures that set a standard of what protocols to use during treatment.^14^ This leads to lack of reproducibility and the inability to compare one study to another.

Another factor that creates the incompatibility between studies is the broad range in the type of assessment used. One review article counted 16 different types of outcome measures used in the studies.^17^

Other factors to consider within given studies include maturation effects, the experience of therapists providing treatment, other medications or therapies participants are receiving, duration, rate, and frequency of treatment, and the caregiving effect.

Looking closely at the review papers of SIT one sees that the authors mostly excluded the 4 papers I excluded above. If one excludes the above papers and only includes randomized control trials (RCTs) that compared SIT to usual care one finds that there are only 2 RCTs that were reviewed by the various papers, Pfeiffer (2011)^9^ which are included in the table above and Schaff (2013)^23^ which did not come up in the PubMed search. These studies however did not include the foreign studies included in this paper’s search.^12, 13^

To clarify the matter, in 2022 researchers set out to conduct a clinical study that removed all the confounding factors mentioned above. Randell (2022)^14^ conducted an RCT with 138 participants, the biggest sample size yet. Additionally, the researchers set a protocol of treatment, conducted the treatment with competent therapists, and assessed patients using a recent standardized sensory disorder assessment together with GAS, an individualized goal assessment. The results of that study showed no significant difference between treatment groups. However, it should be noted that the duration of treatment was only difference between this study and other studies, - twice a week over 10 weeks then twice a month for 3 months.

Future studies should maintain the high standard of randomized clinical trials together with the controls that were mentioned above. Research should vary the protocol used - SIT together with the duration rate and frequency to see if there is any evidence of positive outcomes.

### Other Treatments

As mentioned above in the results, review articles have concluded that SBI therapy has not proven to have significant positive outcomes. The few papers that reviewed AIT, massage therapy and music therapy concluded that either there is no evidence of positive outcomes or that the papers need more confidence in the studies that produced positive outcomes. More research is required to further clarify the outcomes of these therapies.

### Conclusion

Sensory processing disorder in children with ASD has no clear treatment. Sensory integration therapy has shown the most promise of producing positive outcomes. Other therapies have shown little evidence that they produce more results than usual care. Though some studies have shown positive outcomes in SIT many studies have shown no evidence of such. There are many confounding factors within the research and extracting evidence of positive outcomes from a review of the studies is unfeasible. Further research is required to clarify if SIT should become the standard of treatment for these children. Practitioners of patients with ASD who present with sensory processing disorders should consider the outcomes of these studies before referring the patient to therapies mentioned in this paper.

## Data Availability

All data produced in the present work are contained in the manuscript

## References

1. Centers for Disease Control and Prevention. Data & statistics on autism spectrum disorder. Centers for Disease Control and Prevention. Published December 2, 2021. https://www.cdc.gov/ncbddd/autism/data.html

1. Autism Spectrum Disorder. NIMH. www.nimh.nih.gov. https://www.nimh.nih.gov/health/topics/autism-spectrum-disorders-asd#part_145440#:~:text=Overview.%20Autism%20spectrum%20disorder%20%28ASD%29%20is%20a%20developmental

3. Case-Smith J, Weaver LL, Fristad MA. A systematic review of sensory processing interventions for children with autism spectrum disorders. Autism. 2015;19(2):133–148. doi:10.1177/1362361313517762

4. Klintwall L, Holm A, Eriksson M, et al. Sensory abnormalities in autism. A brief report. Res Dev Disabil. 2011;32(2):795–800. doi:10.1016/j.ridd.2010.10.021

5. Robertson CE, Baron-Cohen S. Sensory perception in autism. Nat Rev Neurosci. 2017;18(11):671–684. doi:10.1038/nrn.2017.112

6. Ben-Sasson A, Hen L, Fluss R, Cermak SA, Engel-Yeger B, Gal E. A meta-analysis of sensory modulation symptoms in individuals with autism spectrum disorders. J Autism Dev Disord. 2009;39(1):1–11. doi:10.1007/s10803-008-0593-3

7. Schaaf RC, Case-Smith J. Sensory interventions for children with autism [published correction appears in J Comp Eff Res. 2014 Jul;3(4):446]. J Comp Eff Res. 2014;3(3):225-227. doi:10.2217/cer.14.18

8. Case-Smith J, Bryan T. The effects of occupational therapy with sensory integration emphasis on preschool-age children with autism. Am J Occup Ther. 1999;53(5):489–497. doi:10.5014/ajot.53.5.489

9. Pfeiffer BA, Koenig K, Kinnealey M, Sheppard M, Henderson L. Effectiveness of sensory integration interventions in children with autism spectrum disorders: a pilot study. Am J Occup Ther. 2011;65(1):76–85. doi:10.5014/ajot.2011.09205

10. Iwanaga R, Honda S, Nakane H, Tanaka K, Toeda H, Tanaka G. Pilot study: efficacy of sensory integration therapy for Japanese children with high-functioning autism spectrum disorder. Occup Ther Int. 2014;21(1):4–11. doi:10.1002/oti.1357

11. Karim AEA, Mohammed AH. Effectiveness of sensory integration program in motor skills in children with autism. Egyptian Journal of Medical Human Genetics. 2015: 16(4) 375–380. doi:org/10.1016/j.ejmhg.2014.12.008

12. Kashefimehr B, Kayihan H, Huri M. The Effect of Sensory Integration Therapy on Occupational Performance in Children With Autism. OTJR (Thorofare N J). 2018;38(2):75–83. doi:10.1177/1539449217743456

13. Xu W, Yao J, Liu W. Intervention Effect of Sensory Integration Training on the Behaviors and Quality of Life of Children with Autism. Psychiatr Danub. 2019;31(3):340–346. doi:10.24869/psyd.2019.340

14. Randell E, McNamara R, Delport S, et al. Sensory integration therapy versus usual care for sensory processing difficulties in autism spectrum disorder in children: study protocol for a pragmatic randomised controlled trial. Trials. 2019;20(1):113. Published 2019 Feb 11. doi:10.1186/s13063-019-3205-y

15. Russell L, O’Reilly M, Healy O, Rispoli M, Lyson H, Streusand W, Davis T, Kang S, Sigafoos J, Lancioni G, et al. Sensory integration therapy for autism spectrum disorders: A systematic review. Research in Autism Spectrum Disorders. 2012;6(3)1004-1018. 10.1016/j.rasd.2012.01.006.

16. Case-Smith J, Weaver LL, Fristad MA. A systematic review of sensory processing interventions for children with autism spectrum disorders. Autism. 2015;19(2):133–148. doi:10.1177/1362361313517762

17. Watling R, Hauer S. Effectiveness of Ayres Sensory Integration® and Sensory-Based Interventions for People With Autism Spectrum Disorder: A Systematic Review. Am J Occup Ther. 2015;69(5):6905180030p1-6905180030p12. doi:10.5014/ajot.2015.018051

18. Weitlauf AS, Sathe N, McPheeters ML, Warren ZE. Interventions Targeting Sensory Challenges in Autism Spectrum Disorder: A Systematic Review. Pediatrics. 2017;139(6):e20170347. doi:10.1542/peds.2017-0347

19. Schaaf RC, Dumont RL, Arbesman M, May-Benson TA. Efficacy of Occupational Therapy Using Ayres Sensory Integration^®^: A Systematic Review. Am J Occup Ther. 2018;72(1):7201190010p1-7201190010p10. doi:10.5014/ajot.2018.028431

20. Schoen SA, Lane SJ, Mailloux Z, et al. A systematic review of ayres sensory integration intervention for children with autism. Autism Res. 2019;12(1):6–19. doi:10.1002/aur.2046

21. Case-Smith J, Weaver LL, Fristad MA. A systematic review of sensory processing interventions for children with autism spectrum disorders. Autism. 2015;19(2):133–148. doi:10.1177/1362361313517762

22. Al-Ayadhi L, Al-Drees AM, Al-Arfaj AM. Effectiveness of Auditory Integration Therapy in Autism Spectrum Disorders-Prospective Study. Libertas Academica. 2013. Doi. 10.4137/AUI.S11463

23. Schaaf RC, Case-Smith J. Sensory interventions for children with autism [published correction appears in J Comp Eff Res. 2014 Jul;3(4):446]. J Comp Eff Res. 2014;3(3):225-227. doi:10.2217/cer.14.18

24. Rogers SJ, Ozonoff S. Annotation: what do we know about sensory dysfunction in autism? A critical review of the empirical evidence. J Child Psychol Psychiatry. 2005;46(12):1255–1268. doi:10.1111/j.1469-7610.2005.01431.x

25. Section On Complementary And Integrative Medicine; Council on Children with Disabilities; American Academy of Pediatrics, Zimmer M, Desch L. Sensory integration therapies for children with developmental and behavioral disorders. Pediatrics. 2012;129(6):1186–1189. doi:10.1542/peds.2012-0876

26. Gentil-Gutiérrez A, Cuesta-Gómez JL, Rodríguez-Fernández P, González-Bernal JJ. Implication of the Sensory Environment in Children with Autism Spectrum Disorder: Perspectives from School. Int J Environ Res Public Health. 2021;18(14):7670. Published 2021 Jul 19. doi:10.3390/ijerph18147670

